# Transcriptomics Meta-Analysis Predicts Two Robust Human Biomarkers for Severe Infection with SARS-CoV-2

**DOI:** 10.1101/2022.06.06.22276040

**Authors:** Jeffrey Clancy, Curtis S. Hoffmann, Brett E. Pickett

**Author notes:** Corresponding author: Brett E. Pickett, Corresponding author.

## Abstract

Defining the human factors associated with severe vs mild SARS-CoV-2 infection has become of increasing interest. Mining large numbers of public gene expression datasets is an effective way to identify genes that contribute to a given phenotype. Combining RNA-sequencing data with the associated clinical metadata describing disease severity can enable earlier identification of patients who are at higher risk of developing severe COVID-19 disease. We consequently identified 358 public RNA-seq human transcriptome samples from the Gene Expression Omnibus database that had disease severity metadata. We then subjected these samples to a robust RNA-seq data processing workflow to quantify gene expression in each patient. This process involved using Salmon to map the reads to the reference transcriptomes, edgeR to calculate significant differential expression levels, and gene ontology enrichment using Camera. We then applied a machine learning algorithm to the read counts data to identify features that best differentiated samples based on COVID-19 severity phenotype. Ultimately, we produced a ranked list of genes based on their Gini importance values that includes GIMAP7 and S1PR2, which are associated with immunity and inflammation (respectively). Our results show that these two genes can potentially predict people with severe COVID-19 at up to ∼90% accuracy. We expect that our findings can help contribute to the development of improved prognostics for severe COVID-19.

## Introduction

Human infections with severe Acute Respiratory Syndrome Coronavirus 2 (SARS-CoV-2) has resulted in hundreds of millions of confirmed cases and millions of deaths globally. In addition, countless others have been hospitalized and a subset of the infected population has experienced severe health consequences--particularly those who are elderly, immunocompromised, or have other underlying conditions. The genetic material for this pathogen consists of a monopartite positive-sense single-stranded RNA molecule that is approximately 30 kb in length that contains multiple open reading frames [1]. Since the virus was first detected in late 2019, the scientific community has performed multiple studies to better understand the underlying mechanism(s) of entry and pathogenesis [2–6].

Human pathogenesis studies performed early in the SARS-CoV-2 pandemic showed that the virus induces the interferon response and interleukin-6, as well as other cytokines and chemokines that contribute to COVID-19 [7–9]. Interestingly, multiple studies have shown that the majority of infections are either mild or asymptomatic [10–13]. The large diversity in the human response to infection, combined with large numbers of infections, contributed to strained hospital capacity [14–16]. Various factors contribute to these observed differences in disease severity, and the demand for robust biomarkers associated with COVID-19 disease severity has continually grown. Some studies have identified associations between acute infection and the host response [17,18]. Other studies have evaluated associations between disease severity and aspects of the adaptive immune system [19–23] and quantified viral RNA [24–29]. A recent study has used neural networks to predict patient survival outcomes with high accuracy [30], which can be useful when whole transcriptome data are available. However, to our knowledge, a meta-analysis on transcriptional biomarkers associated with mild versus severe infection has not been previously reported.

The aim of the current study is to perform a meta-analysis of existing human transcriptomics data from collected blood samples to predict transcriptional prognostic markers. Since presence of the virus is possible with existing diagnostics, such markers of disease severity could then contribute to making informed decisions concerning the care of infected patients who seek treatment at the hospital.

## Methods

### Identification of Relevant Datasets

An established process that combines automated and manual methods was used to identify and analyze the samples from published SARS-CoV-2 human transcriptomics studies with metadata specifying the severity of infection in May, 2021. Samples from patients having either “mild” or “asymptomatic” infections were manually labeled by one reviewer as “mild”, while those recorded as “hospitalized”, “ICU”, or “death” were labeled as “severe”. Records (GSE152418 (PBMC), GSE157103 (whole blood), GSE166424 (whole blood)) in the Gene Expression Omnibus (GEO) database [31–33], within the National Center for Biotechnology Information (NCBI), were queried and manually reviewed for relevance (Figure 1). Specifically, studies were selected based on the presence of three predefined criteria: 1) the host organism was human, 2) the data were generated as part of a RNA-sequencing experiment, and 3) the study included samples collected during acute SARS-CoV-2 infection together with disease severity metadata. In total, we identified and processed 358 relevant samples across three independent RNA-sequencing studies.

**Figure 1:**
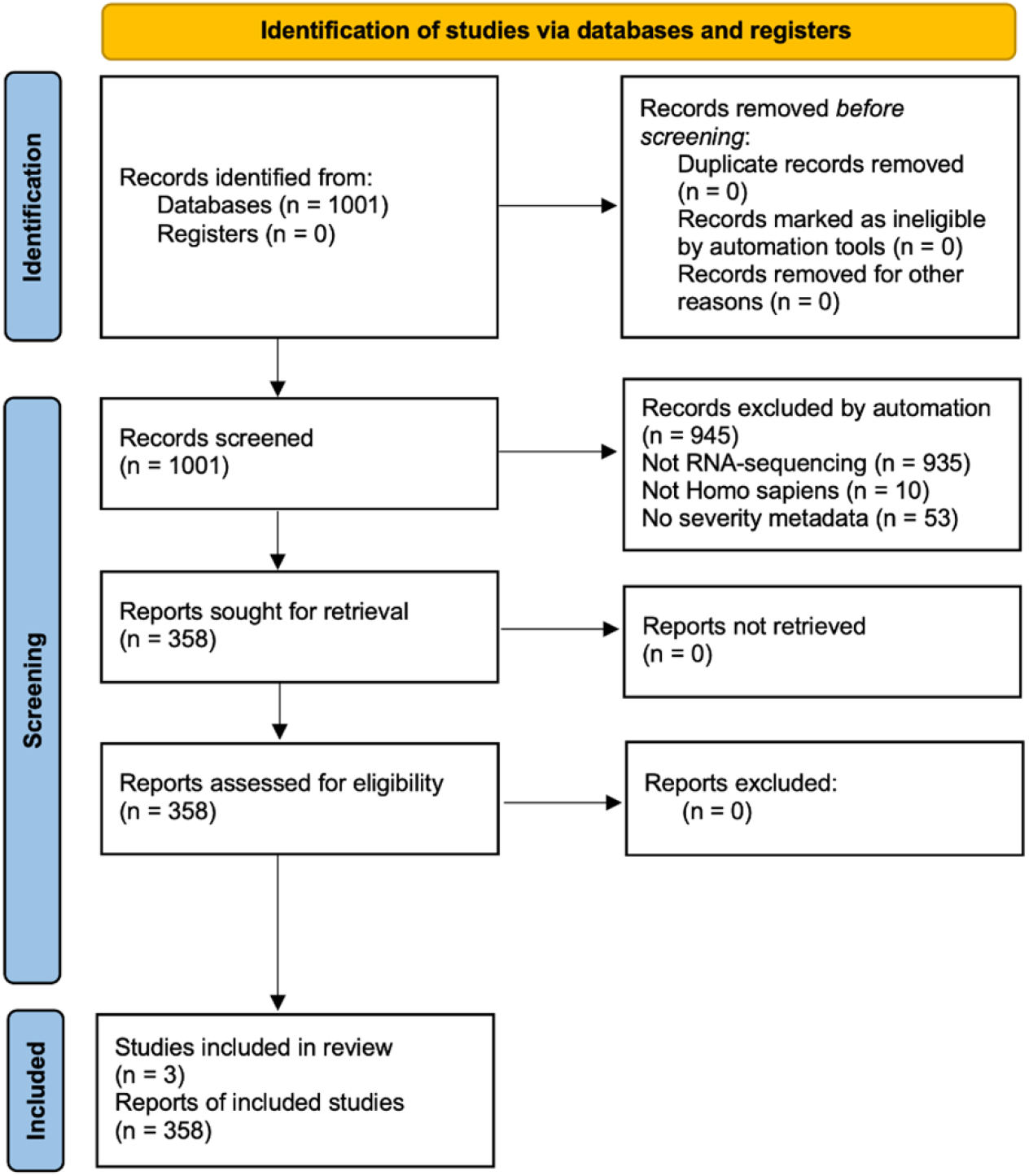
Flow diagram to visualize the process used to filter relevant public studies, samples, and files. **PRISMA 2020 flow diagram for new systematic reviews which included searches of databases and registers only** *From:* Page MJ, McKenzie JE, Bossuyt PM, Boutron I, Hoffmann TC, Mulrow CD, et al. The PRISMA 2020 statement: an updated guideline for reporting systematic reviews. BMJ 2021;372:n71. doi: 10.1136/bmj.n71 For more information, visit: http://www.prisma-statement.org/

### Data Pre-Processing

The fastq files containing the RNA-sequencing data were obtained from the Sequence Read Archive (SRA) at NCBI [34] using the sratools software. These files were divided into “case” and “control” categories based on whether they had “severe” or “mild” disease. The Automated Reproducible MOdular Workflow for Preprocessing and Differential Analysis of RNA-seq Data (ARMOR) was then used to preprocess and analyze the RNA-seq data [35]. Briefly, this ARMOR workflow uses the python-based snakemake workflow language [36] to perform steps including: trimming of sequencing adapters and low-quality regions from the originally-generated RNA-sequencing reads with TrimGalore (https://www.bioinformatics.babraham.ac.uk/projects/trim_galore/), calculate quality control metrics with FastQC (www.bioinformatics.babraham.ac.uk/projects/fastqc/), as well as map and quantify reads to the human GRCh38 transcriptome with Salmon [37]. Differential gene expression was calculated from the read counts with edgeR [38]. Gene ontology terms were calculated from the list of significant gene identifiers produced by edgeR by using the DAVID resource [39].

### Machine Learning

The salmon counts for each gene in each GEO sample identifier were compiled into a table. The counts for each sample were then normalized using a z-score transformation. These samples into test and training sets, with 70% of samples assigned to a training set and the remaining 30% of samples assigned to a test set. A random forest classification method (using the R randomforest package) was then trained and used to identify the genes that were most useful in classifying samples as coming from patients with either “severe” or “mild” COVID-19 disease. This random forest algorithm calculated the Gini impurity values for each feature, which were then sorted to rank the importance of genes based on the original training set. This process was repeated for subsets of the best-scoring features to quantify their accuracy, specificity, and sensitivity.

## Results

### Transcriptomics Meta-Analysis Identifies Significant Genes, Enriched Terms, and Signaling Pathways

We began by identifying publicly available RNA-sequencing data generated from either whole blood samples or peripheral blood mononuclear cells (PBMCs) that had been previously collected from patients infected with SARS-CoV-2 and had associated disease severity metadata. We then assigned these samples to either “high severity” or “low severity”. We processed these RNA-seq files using an automated computational workflow that performed quality control, trimmed reads, mapped them to the human transcriptome, and calculated significant differentially-expressed genes. We then used these genes to identify enriched Gene Ontology (GO) terms.

Overall, we identified 8176 significant differentially expressed genes after applying a multiple hypothesis correction with log_2_ fold-change values ranging from -4.2 to 3.78 (Figure 2). We found that the most significant differentially expressed genes included ASPH, C5orf30, DGKH, SLC26A6. We then subjected this list of significant DEGs to Gene Ontology enrichment. This analysis produced 90 significant GO terms including immune response, apoptosis, and I-kappaB kinase/NF-kappaB signaling (Table 1). It is possible that a subset of these results may contain bias due to the heterogeneity of the samples. However, we expect that the robust statistical analysis of these samples minimized such effects.

**Table 1:** The most significant Gene Ontology Terms generated from DAVID.

| GO Term | Count | % | P-Value | Corrected P-Value |
| --- | --- | --- | --- | --- |
| protein binding | 5518 | 69.4 | 3.10E-105 | 9.70E-102 |
| RNA binding | 731 | 9.2 | 2.80E-21 | 4.30E-18 |
| identical protein binding | 802 | 10.1 | 2.20E-14 | 2.30E-11 |
| metal ion binding | 1188 | 15 | 2.90E-13 | 2.20E-10 |
| regulation of catalytic activity | 215 | 2.7 | 7.90E-12 | 5.10E-08 |
| apoptotic process | 296 | 3.7 | 1.10E-11 | 5.10E-08 |
| DNA repair | 158 | 2 | 2.60E-11 | 8.00E-08 |
| catalytic activity | 89 | 1.1 | 6.40E-11 | 3.90E-08 |
| regulation of transcription, DNA-templated | 456 | 5.7 | 7.60E-11 | 1.80E-07 |
| positive regulation of I-kappaB kinase/NF-kappaB signaling | 115 | 1.4 | 2.00E-10 | 3.60E-07 |
| ATP binding | 710 | 8.9 | 5.00E-10 | 2.60E-07 |
| protein phosphorylation | 255 | 3.2 | 1.30E-09 | 2.00E-06 |
| ubiquitin protein<br>ligase binding | 168 | 2.1 | 5.40E-09 | 2.40E-06 |
| protein transport | 221 | 2.8 | 7.50E-09 | 1.00E-05 |
| protein<br>ubiquitination | 244 | 3.1 | 9.30E-09 | 1.10E-05 |
| innate immune<br>response | 281 | 3.5 | 1.30E-08 | 1.40E-05 |
| adaptive immune<br>response | 217 | 2.7 | 1.60E-08 | 1.50E-05 |
| ubiquitin-protein<br>transferase activity | 141 | 1.8 | 1.80E-08 | 7.10E-06 |

**Figure 2.**
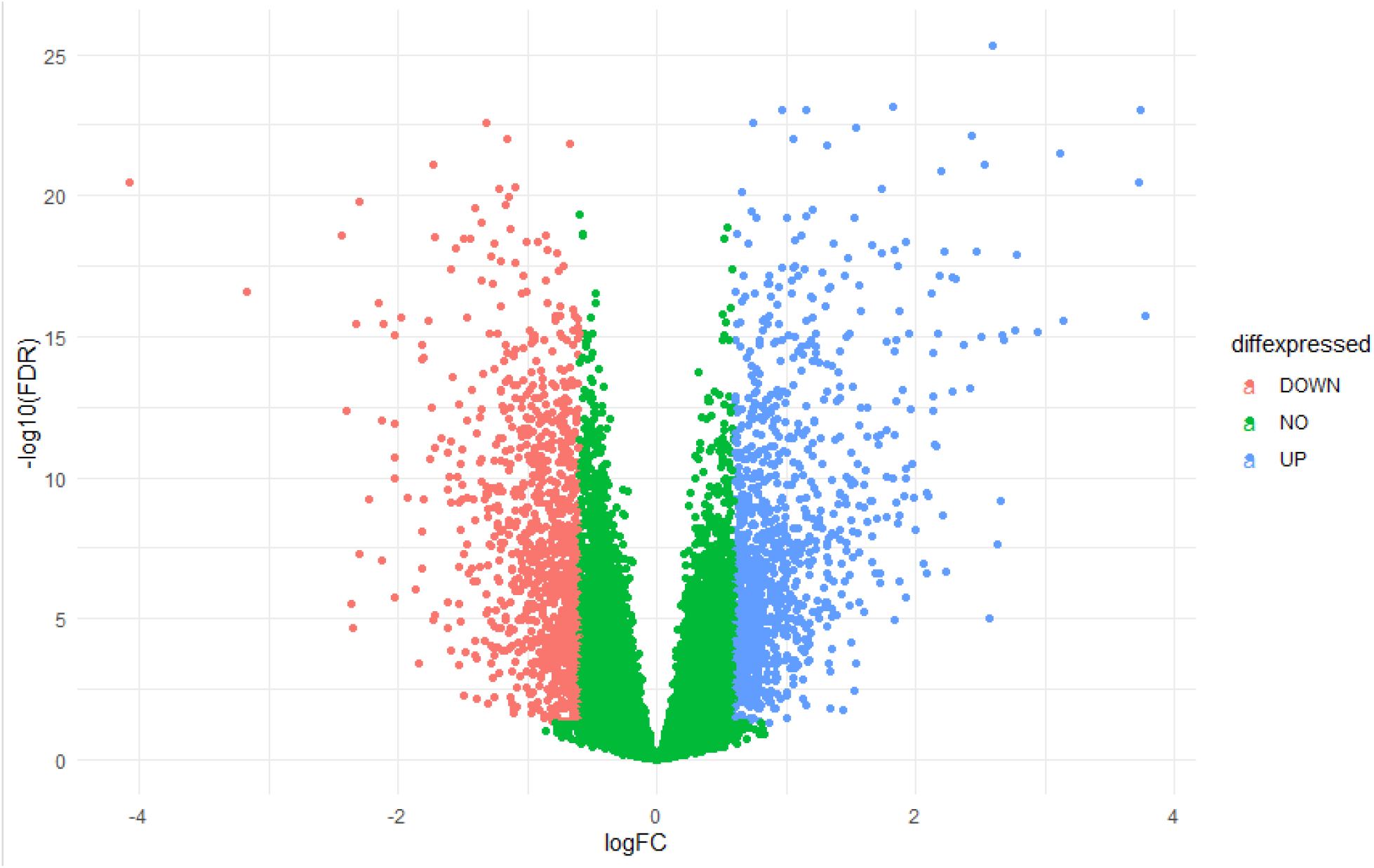
Volcano plot of all differentially expressed genes in severe vs. mild human infection with SARS-CoV-2. Genes that are up or down regulated from blood samples collected from patients having severe symptoms or mild symptoms during infection with SARS-CoV-2. Genes showing statistically significant up-regulation (blue), down-regulated (red), or no significant change (green). X-axis shows the log_2_ fold-change values while the y-axis displays false-discovery rate-adjusted p-values to account for multiple hypothesis testing.

We then used the signaling pathway impact analysis algorithm to calculate which intracellular signaling pathways were best represented by the list of DEGs. This analysis identified nine pathways that were significantly affected by severe COVID-19 (Table 2). We observed that five of these significant pathways dealt directly with T-cell receptor (TCR) signaling, while a sixth described a Zap70 immunological synapse. Interestingly, all six of these immune-related pathways were predicted to be inhibited during severe COVID-19.

**Table 2:**
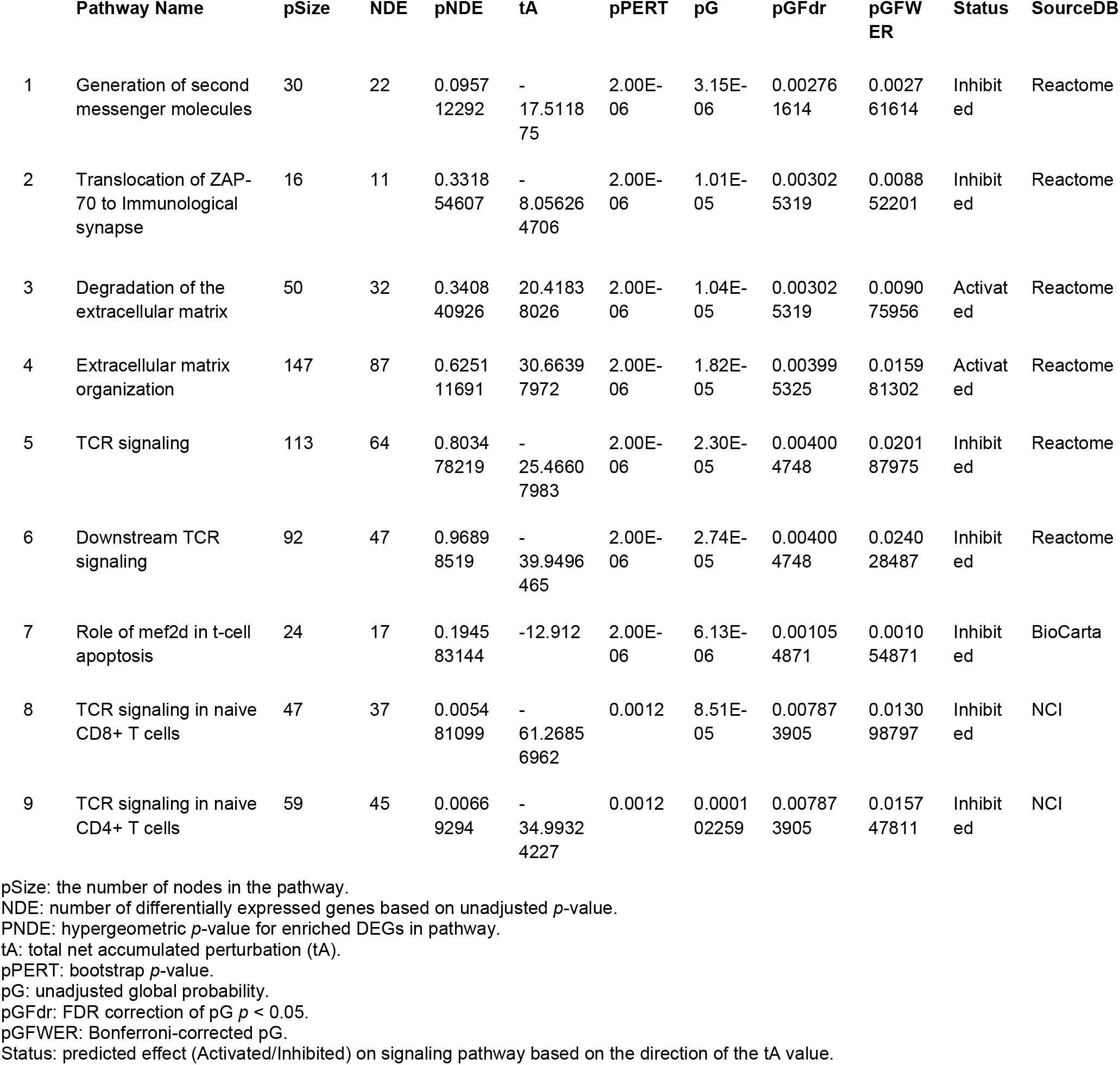
Significant Intracellular Signaling Pathways During Severe COVID-19.

|  | Pathway Name | pSize | NDE | pNDE | tA | pPERT | pG | pGFdr | pGFWER | Status | SourceDB |
| --- | --- | --- | --- | --- | --- | --- | --- | --- | --- | --- | --- |
| 1 | Generation of second messenger molecules | 30 | 22 | 0.0957<br>12292 | -<br>17.5118<br>75 | 2.00E-06 | 3.15E-06 | 0.00276<br>1614 | 0.0027<br>61614 | Inhibit<br>ed | Reactome |
| 2 | Translocation of ZAP-70 to Immunological synapse | 16 | 11 | 0.3318<br>54607 | -<br>8.05626<br>4706 | 2.00E-06 | 1.01E-05 | 0.00302<br>5319 | 0.0088<br>52201 | Inhibit<br>ed | Reactome |
| 3 | Degradation of the extracellular matrix | 50 | 32 | 0.3408<br>40926 | 20.4183<br>8026 | 2.00E-06 | 1.04E-05 | 0.00302<br>5319 | 0.0090<br>75956 | Activat<br>ed | Reactome |
| 4 | Extracellular matrix organization | 147 | 87 | 0.6251<br>11691 | 30.6639<br>7972 | 2.00E-06 | 1.82E-05 | 0.00399<br>5325 | 0.0159<br>81302 | Activat<br>ed | Reactome |
| 5 | TCR signaling | 113 | 64 | 0.8034<br>78219 | -<br>25.4660<br>7983 | 2.00E-06 | 2.30E-05 | 0.00400<br>4748 | 0.0201<br>87975 | Inhibit<br>ed | Reactome |
| 6 | Downstream TCR signaling | 92 | 47 | 0.9689<br>8519 | -<br>39.9496<br>465 | 2.00E-06 | 2.74E-05 | 0.00400<br>4748 | 0.0240<br>28487 | Inhibit<br>ed | Reactome |
| 7 | Role of mef2d in t-cell apoptosis | 24 | 17 | 0.1945<br>83144 | -12.912 | 2.00E-06 | 6.13E-06 | 0.00105<br>4871 | 0.0010<br>54871 | Inhibit<br>ed | BioCarta |
| 8 | TCR signaling in naive CD8+ T cells | 47 | 37 | 0.0054<br>81099 | -<br>61.2685<br>6962 | 0.0012 | 8.51E-<br>05 | 0.00787<br>3905 | 0.0130<br>98797 | Inhibit<br>ed | NCI |
| 9 | TCR signaling in naive CD4+ T cells | 59 | 45 | 0.0066<br>9294 | -<br>34.9932<br>4227 | 0.0012 | 0.0001<br>02259 | 0.00787<br>3905 | 0.0157<br>47811 | Inhibit<br>ed | NCI |

pSize: the number of nodes in the pathway.
NDE: number of differentially expressed genes based on unadjusted *p*-value.
pNDE: hypergeometric *p*-value for enriched DEGs in pathway.
tA: total net accumulated perturbation (tA).
pPERT: bootstrap *p*-value.
pG: unadjusted global probability.
pGFdr: FDR correction of pG $p < 0.05$ .
pGFWER: Bonferroni-corrected pG.
Status: predicted effect (Activated/Inhibited) on signaling pathway based on the direction of the tA value.

### Machine Learning Identifies GIMAP7 and S1PR2 as Robust Biomarkers

Prior to predicting relevant biomarkers, we first wanted to confirm whether the intracellular transcriptional response in blood was strongly associated with the disease severity metadata that was recorded with each record. We consequently constructed a table with all transcripts from each gene represented as columns and the read mapping data represented as rows. We then used this table to generate a receiver-operator characteristic (ROC) curve (Figure 3). In this case, the area under the curve (AUC) represents the percent specificity and sensitivity for the host transcriptomic data to predict disease severity. We calculated the AUC of the curve across all transcripts to be 96.6%, which indicates that the host transcriptional response strongly contributes to disease severity.

**Figure 3.**
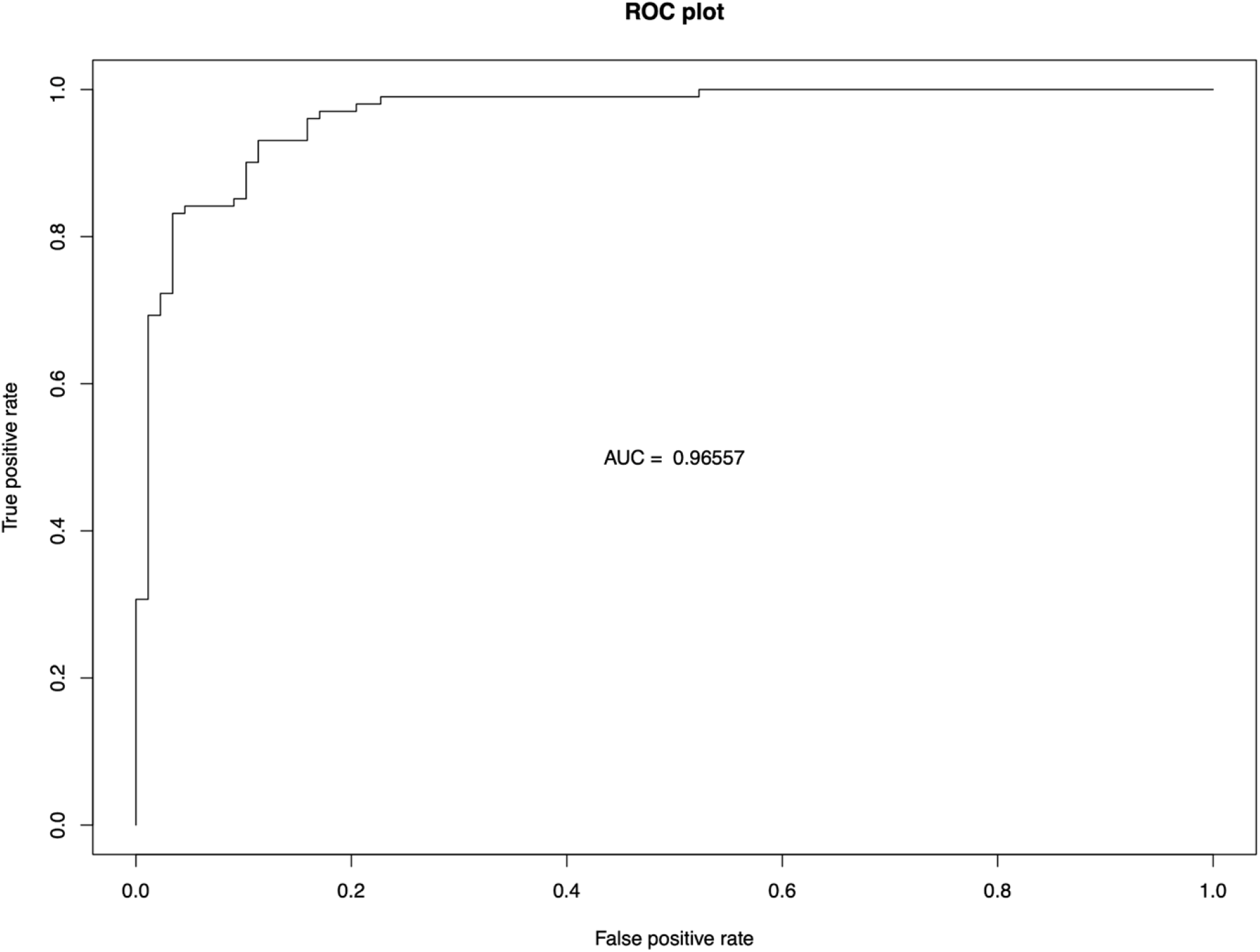
Receiver-operator characteristic (ROC) curve constructed from all expressed genes in severe vs. mild human infection with SARS-CoV-2. Constructing a ROC curve from all RNA-sequencing read quantification values achieved an area-under-the-curve (AUC) value of greater than 96%.

We then subjected the same tabular data to a machine learning algorithm to enable us to predict the features (e.g. expressed genes) that were most associated with the disease severity phenotype (Table 3). We ranked our random forest output by descending order of the Mean Decrease in Gini Impurity value, which is a measure of entropy. Transcripts from genes with larger Gini Impurity values represented those that could be used to most accurately predict the recorded disease phenotype.

**Table 3:**
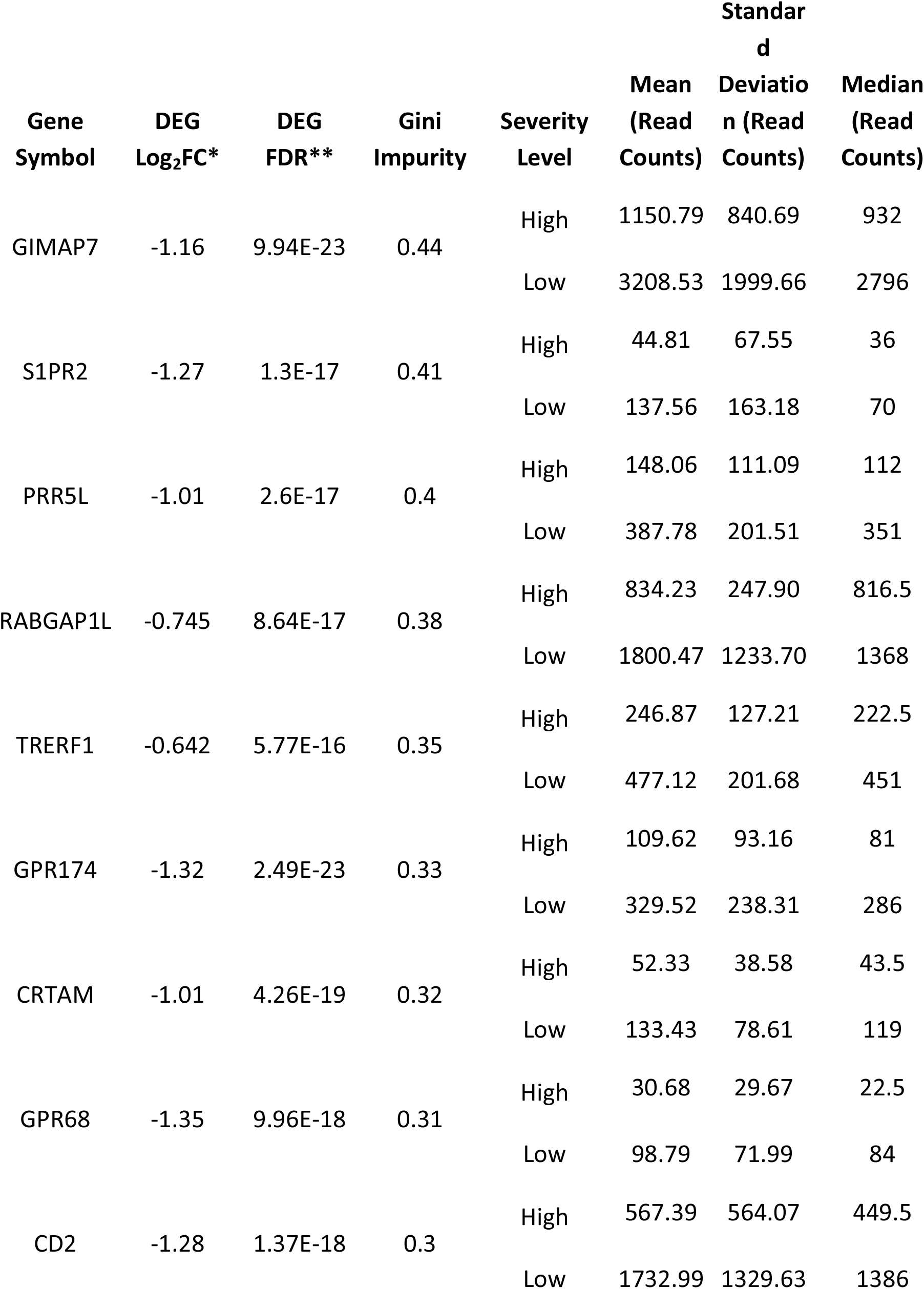
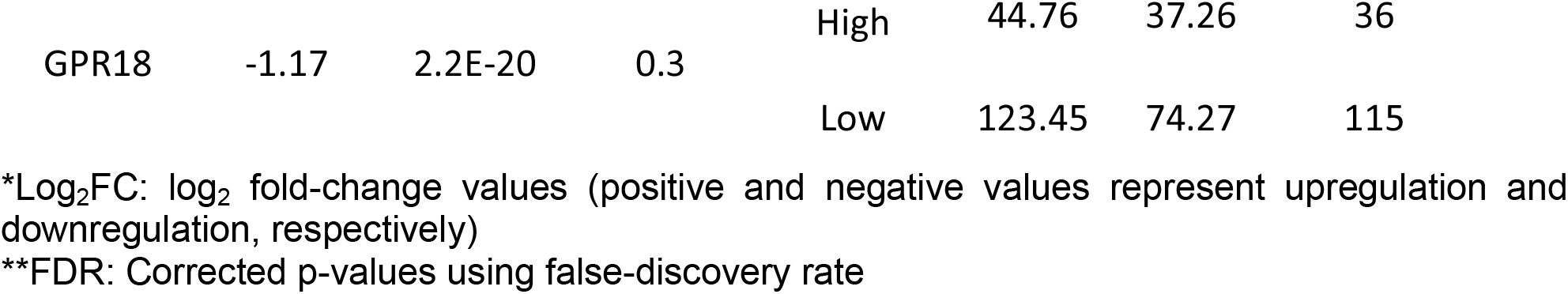
List of top 10 expressed genes that best predicted SARS-CoV-2 disease severity in human blood cells.

We next wanted to reduce the selected markers to the smallest number that would provide the best performance. To do so, we generated a ROC curve for the six expressed genes having the highest Gini Impurity values and calculated its AUC to be 94.3%. We repeated this process for only the top two expressed genes (GIMAP7 and S1PR2) and only the top expressed gene (GIMAP7). This analysis quantified the AUC to be 89.8% for the two combined genes and 84.4% for only the top gene. Specifically, the mean and median read counts for each of these two genes were approximately three times higher in the samples with low disease severity than in the samples with high disease severity.

## Discussion

Previous work has shown that combining multiple datasets in meta-analyses can augment the signal-to-noise ratio in order to gain new biological and mechanistic insight(s) [9,40–43]. As such, the goal of our study was to predict prognostic markers of SARS-CoV-2 disease severity through a transcriptomics meta-analysis of human blood samples. We found statistically significant differentially expressed genes, enriched Gene Ontology terms, modulated signaling pathways, and a ranked list of biomarkers that could potentially be combined to predict which patients are at risk of severe disease.

At least one prior study has generated single-cell RNA-sequencing data to better understand the host response to SARS-CoV-2 infection [44]. Their findings showed adaptive immune components play a role in disease severity. Interestingly, our data confirm results from some prior experiments that show certain aspects of the T-cell response may be downregulated during SARS-CoV-2 infection [45–47]. A modified distribution of the ZAP70 kinase on the plasma membrane of T-cells contributes to the signal transduction and amplification of the TCR [48]. Interestingly, CD3/ZAP70 protein has been shown to interact with TREM-2 in the T-cells of patients infected with SARS-CoV-2 [49]. The ASPH and C5orf30 genes, which were identified as significant DEGs in our study, have been identified previously as markers of severe infection [50].

These prior studies support our finding that GIMAP7 could be important in severity given its presence on the surface of T-lymphocytes [51]. In the time since GIMAP7 was initially identified [52], it has subsequently been shown to be a potential marker for various cancer types, which further supports its immune-related role [53–56]. Although the underlying mechanism for this protein is yet to be characterized, we are not surprised by its role in other immune-related diseases or in SARS-CoV-2 disease severity. Information about the S1PR2 gene is more sparse, although it appears to have a role in inflammation and other immune-related functions [57–61]. Specifically, S1PR2 on T-cells recruits lymphocytes to damaged tissues and may contribute to the recirculation of cells in the adaptive immune system to the lymphatic system [62].

The GIMAP7 gene, together with a subset of the enriched GO terms, had been found in an earlier study on transcriptional biomarkers for SARS-CoV-2 [63]; however, it was not highly ranked. This is logical since DEGs are often identified as biomarkers. In addition, as the sample size increases among a more diverse patient population, we expect that the genes most capable of differentiating disease severity should become more accurate.

It is important to note that these samples were taken while the early variants were circulating. As such, further testing would be required to confirm whether these biomarkers are still consistent predictors of infection severity in patients infected with more recent variants. We anticipate that qRT-PCR (quantitative Reverse Transcriptase Polymerase Chain reaction), and possibly flow cytometry could be used to quantify these biomarkers in relevant samples. Additional experiments are needed to confirm whether these findings can be replicated in samples across different age and/or risk groups.

These results present a potential predictor for the severity levels of patients infected with SARS-CoV-2. Considering the diversity of reactions that patients have to the virus, incorporating these biomarkers as additional data points to assess patient risk of severe disease could be pivotal in augmenting both personal and public health decision making processes [64–66]. This is especially relevant when resources may be limited and priority must be given to those with the greatest risk of severe infection. These results in particular are useful because they illustrate both up and down regulation association that best differentiate each patient in a population, rather than just identifying genes with statistically significant changes across the populations being compared. This approach allows us to detect the directionality of biomarkers such as GIMAP7 and S1PR2 that were not as highly ranked by edgeR.

In conclusion, the findings from this study could contribute to ongoing efforts relating to triaging patients when hospitals are approaching their capacity limit. We envision that creating an assay to quantify the presence of a subset of the features identified in this study could be useful in identifying patients who are at higher risk of developing severe disease. The results of such a prognostic assay could contribute to triage efforts and to treatment decisions being made in the clinic.

## Data Availability

Publicly available RNA-seq fastq files were obtained from the NCBI Gene Expression Omnibus. The relevant studies include: GSE152418, GSE157103, and GSE166424.

https://www.ncbi.nlm.nih.gov/geo/query/acc.cgi?acc=GSE152418

https://www.ncbi.nlm.nih.gov/geo/query/acc.cgi?acc=GSE157103

https://www.ncbi.nlm.nih.gov/geo/query/acc.cgi?acc=GSE166424

## Acknowledgements

We are grateful to the BYU Office of Research Computing for providing the computational resources needed to complete this study. We also thank the original clinicials, patients, and others who provided the RNA-sequencing data.

## References

1. Kim D, Lee J-Y, Yang J-S, Kim JW, Kim VN, Chang H. The Architecture of SARS-CoV-2 Transcriptome. Cell. 2020;181: 914–921.e10.

2. Yan R, Zhang Y, Li Y, Xia L, Guo Y, Zhou Q. Structural basis for the recognition of SARS-CoV-2 by full-length human ACE2. Science. 2020;367: 1444–1448.

3. Evans JP, Liu S-L. Role of host factors in SARS-CoV-2 entry. J Biol Chem. 2021;297: 100847.

4. Harrison AG, Lin T, Wang P. Mechanisms of SARS-CoV-2 Transmission and Pathogenesis. Trends Immunol. 2020;41: 1100–1115.

5. Cevik M, Kuppalli K, Kindrachuk J, Peiris M. Virology, transmission, and pathogenesis of SARS-CoV-2. BMJ. 2020;371: m3862.

6. Lamers MM, Haagmans BL. SARS-CoV-2 pathogenesis. Nat Rev Microbiol. 2022;20: 270–284.

7. Blanco-Melo D, Nilsson-Payant BE, Liu W-C, Uhl S, Hoagland D, Møller R, et al. Imbalanced Host Response to SARS-CoV-2 Drives Development of COVID-19. Cell. 2020;181: 1036–1045.e9.

8. Giamarellos-Bourboulis EJ, Netea MG, Rovina N, Akinosoglou K, Antoniadou A, Antonakos N, et al. Complex Immune Dysregulation in COVID-19 Patients with Severe Respiratory Failure. Cell Host Microbe. 2020;27: 992–1000.e3.

9. Ferrarini MG, Lal A, Rebollo R, Gruber AJ, Guarracino A, Gonzalez IM, et al. Genome-wide bioinformatic analyses predict key host and viral factors in SARS-CoV-2 pathogenesis. Commun Biol. 2021;4: 590.

10. Asadi S, Bouvier N, Wexler AS, Ristenpart WD. The coronavirus pandemic and aerosols: Does COVID-19 transmit via expiratory particles? Aerosol Sci Technol. 2020;0: 1–4.

11. Koopmans M, Haagmans B. Assessing the extent of SARS-CoV-2 circulation through serological studies. Nature medicine. 2020. pp. 1171–1172.

12. Syangtan G, Bista S, Dawadi P, Rayamajhee B, Shrestha LB, Tuladhar R, et al. Asymptomatic SARS-CoV-2 Carriers: A Systematic Review and Meta-Analysis. Front Public Health. 2020;8: 587374.

13. Salzberger B, Buder F, Lampl B, Ehrenstein B, Hitzenbichler F, Holzmann T, et al. Epidemiology of SARS-CoV-2. Infection. 2021;49: 233–239.

14. Twohig KA, Nyberg T, Zaidi A, Thelwall S, Sinnathamby MA, Aliabadi S, et al. Hospital admission and emergency care attendance risk for SARS-CoV-2 delta (B.1.617.2) compared with alpha (B.1.1.7) variants of concern: a cohort study. Lancet Infect Dis. 2022;22: 35–42.

15. Sheikh A, McMenamin J, Taylor B, Robertson C, Public Health Scotland and the EAVE II Collaborators. SARS-CoV-2 Delta VOC in Scotland: demographics, risk of hospital admission, and vaccine effectiveness. Lancet. 2021;397: 2461–2462.

16. Finelli L, Gupta V, Petigara T, Yu K, Bauer KA, Puzniak LA. Mortality Among US Patients Hospitalized With SARS-CoV-2 Infection in 2020. JAMA Netw Open. 2021;4: e216556.

17. Gupta RK, Rosenheim J, Bell LC, Chandran A, Guerra-Assuncao JA, Pollara G, et al. Blood transcriptional biomarkers of acute viral infection for detection of pre-symptomatic SARS-CoV-2 infection: a nested, case-control diagnostic accuracy study. Lancet Microbe. 2021;2: e508–e517.

18. Myhre PL, Prebensen C, Jonassen CM, Berdal JE, Omland T. SARS-CoV-2 Viremia is Associated With Inflammatory, But Not Cardiovascular Biomarkers, in Patients Hospitalized for COVID-19. J Am Heart Assoc. 2021;10: e019756.

19. Phipps WS, SoRelle JA, Li Q-Z, Mahimainathan L, Araj E, Markantonis J, et al. SARS-CoV-2 Antibody Responses Do Not Predict COVID-19 Disease Severity. Am J Clin Pathol. 2020;154: 459–465.

20. Singh K, Mittal S, Gollapudi S, Butzmann A, Kumar J, Ohgami RS. A meta-analysis of SARS-CoV-2 patients identifies the combinatorial significance of D-dimer, C-reactive protein, lymphocyte, and neutrophil values as a predictor of disease severity. Int J Lab Hematol. 2021;43: 324–328.

21. Garcia-Beltran WF, Lam EC, Astudillo MG, Yang D, Miller TE, Feldman J, et al. COVID-19-neutralizing antibodies predict disease severity and survival. Cell. 2021;184: 476–488.e11.

22. Neidleman J, Luo X, George AF, McGregor M, Yang J, Yun C, et al. Distinctive features of SARS-CoV-2-specific T cells predict recovery from severe COVID-19. Cell Rep. 2021;36: 109414.

23. Agwa SHA, Elghazaly H, Meteini MSE, Shawky SM, Ali M, Abd Elsamee AM, et al. In Silico Identification and Clinical Validation of a Novel Long Non-Coding RNA/mRNA/miRNA Molecular Network for Potential Biomarkers for Discriminating SARS CoV-2 Infection Severity. Cells. 2021;10. doi:10.3390/cells10113098

24. Le Borgne P, Solis M, Severac F, Merdji H, Ruch Y, Alamé Intern K, et al. SARS-CoV-2 viral load in nasopharyngeal swabs in the emergency department does not predict COVID-19 severity and mortality. Acad Emerg Med. 2021;28: 306–313.

25. Ram-Mohan N, Kim D, Zudock EJ, Hashemi MM, Tjandra KC, Rogers AJ, et al. SARS-CoV-2 RNAemia Predicts Clinical Deterioration and Extrapulmonary Complications from COVID-19. Clin Infect Dis. 2022;74: 218–226.

26. Olea B, Albert E, Torres I, Gozalbo-Rovira R, Carbonell N, Ferreres J, et al. Lower respiratory tract and plasma SARS-CoV-2 RNA load in critically ill adult COVID-19 patients: Relationship with biomarkers of disease severity. The Journal of infection. 2021. pp. 381–412.

27. Wong A, Lin ZQ, Wang L, Chung AG, Shen B, Abbasi A, et al. Towards computer-aided severity assessment via deep neural networks for geographic and opacity extent scoring of SARS-CoV-2 chest X-rays. Sci Rep. 2021;11: 9315.

28. Ghodake GS, Shinde SK, Kadam AA, Saratale RG, Saratale GD, Syed A, et al. Biological characteristics and biomarkers of novel SARS-CoV-2 facilitated rapid development and implementation of diagnostic tools and surveillance measures. Biosens Bioelectron. 2021;177: 112969.

29. Zhang L, Guo H. Biomarkers of COVID-19 and technologies to combat SARS-CoV-2. Adv Biomark Sci Technol. 2020;2: 1–23.

30. Rasmy L, Nigo M, Kannadath BS, Xie Z, Mao B, Patel K, et al. Recurrent neural network models (CovRNN) for predicting outcomes of patients with COVID-19 on admission to hospital: model development and validation using electronic health record data. Lancet Digit Health. 2022. doi:10.1016/S2589-7500(22)00049-8

31. Edgar R, Domrachev M, Lash AE. Gene Expression Omnibus: NCBI gene expression and hybridization array data repository. Nucleic Acids Res. 2002;30: 207–210.

32. Arunachalam PS, Wimmers F, Mok CKP, Perera Rapm, Scott M, Hagan T, et al. Systems biological assessment of immunity to mild versus severe COVID-19 infection in humans. Science. 2020;369: 1210–1220.

33. Overmyer KA, Shishkova E, Miller IJ, Balnis J, Bernstein MN, Peters-Clarke TM, et al. Large-Scale Multi-omic Analysis of COVID-19 Severity. Cell Syst. 2021;12: 23–40.e7.

34. Kodama Y, Shumway M, Leinonen R, International Nucleotide Sequence Database Collaboration. The Sequence Read Archive: explosive growth of sequencing data. Nucleic Acids Res. 2012;40: D54–6.

35. Orjuela S, Huang R, Hembach KM, Robinson MD, Soneson C. ARMOR: An utomated eproducible dular Workflow for Preprocessing and Differential Analysis of NA-seq Data. G3. 2019;9: 2089–2096.

36. Köster J, Rahmann S. Snakemake--a scalable bioinformatics workflow engine. Bioinformatics. 2012;28: 2520–2522.

37. Patro R, Duggal G, Love MI, Irizarry RA, Kingsford C. Salmon provides fast and bias-aware quantification of transcript expression. Nat Methods. 2017;14: 417–419.

38. Robinson MD, McCarthy DJ, Smyth GK. edgeR: a Bioconductor package for differential expression analysis of digital gene expression data. Bioinformatics. 2010;26: 139–140.

39. Sherman BT, Hao M, Qiu J, Jiao X, Baseler MW, Lane HC, et al. DAVID: a web server for functional enrichment analysis and functional annotation of gene lists (2021 update). Nucleic Acids Res. 2022. doi:10.1093/nar/gkac194

40. Cevik M, Tate M, Lloyd O, Maraolo AE, Schafers J, Ho A. SARS-CoV-2, SARS-CoV, and MERS-CoV viral load dynamics, duration of viral shedding, and infectiousness: a systematic review and meta-analysis. Lancet Microbe. 2021;2: e13–e22.

41. Rostami A, Sepidarkish M, Leeflang MMG, Riahi SM, Nourollahpour Shiadeh M, Esfandyari S, et al. SARS-CoV-2 seroprevalence worldwide: a systematic review and meta-analysis. Clin Microbiol Infect. 2021;27: 331–340.

42. Buitrago-Garcia D, Egli-Gany D, Counotte MJ, Hossmann S, Imeri H, Ipekci AM, et al. Occurrence and transmission potential of asymptomatic and presymptomatic SARS-CoV-2 infections: A living systematic review and meta-analysis. PLoS Med. 2020;17: e1003346.

43. Scott TM, Jensen S, Pickett BE. A signaling pathway-driven bioinformatics pipeline for predicting therapeutics against emerging infectious diseases. F1000Res. 2021;10: 330.

44. Zhang F, Gan R, Zhen Z, Hu X, Li X, Zhou F, et al. Adaptive immune responses to SARS-CoV-2 infection in severe versus mild individuals. Signal Transduct Target Ther. 2020;5: 156.

45. Wellington D, Yin Z, Kessler BM, Dong T. Immunodominance complexity: lessons yet to be learned from dominant T cell responses to SARS-COV-2. Curr Opin Virol. 2021;50: 183–191.

46. Beckmann ND, Comella PH, Cheng E, Lepow L, Beckmann AG, Tyler SR, et al. Downregulation of exhausted cytotoxic T cells in gene expression networks of multisystem inflammatory syndrome in children. Nat Commun. 2021;12: 4854.

47. Park MD. Immune evasion via SARS-CoV-2 ORF8 protein? Nat Rev Immunol. 2020;20: 408.

48. Tewari R, Shayahati B, Fan Y, Akimzhanov AM. T cell receptor-dependent S-acylation of ZAP-70 controls activation of T cells. J Biol Chem. 2021;296: 100311.

49. Wu Y, Wang M, Yin H, Ming S, Li X, Jiang G, et al. TREM-2 is a sensor and activator of T cell response in SARS-CoV-2 infection. Sci Adv. 2021;7: eabi6802.

50. Shaath H, Alajez NM. Identification of PBMC-based molecular signature associational with COVID-19 disease severity. Heliyon. 2021;7: e06866.

51. Filén S, Lahesmaa R. GIMAP Proteins in T-Lymphocytes. J Signal Transduct. 2010;2010: 268589.

52. Krücken J, Schroetel RMU, Müller IU, Saïdani N, Marinovski P, Benten WPM, et al. Comparative analysis of the human gimap gene cluster encoding a novel GTPase family. Gene. 2004;341: 291–304.

53. Qin Y, Liu H, Huang X, Huang L, Liao L, Li J, et al. GIMAP7 as a potential predictive marker for pan-cancer prognosis and immunotherapy efficacy. J Inflamm Res. 2022;15: 1047–1061.

54. Song Y, Pan Y, Liu J. The relevance between the immune response-related gene module and clinical traits in head and neck squamous cell carcinoma. Cancer Manag Res. 2019;11: 7455–7472.

55. Zhang J, Wang L, Xu X, Li X, Guan W, Meng T, et al. Transcriptome-Based Network Analysis Unveils Eight Immune-Related Genes as Molecular Signatures in the Immunomodulatory Subtype of Triple-Negative Breast Cancer. Front Oncol. 2020;10: 1787.

56. Xi Y, Jing Z, Haihong L, Yizhen J, Weili G, Shuwen H. Analysis of T lymphocyte-related biomarkers in pancreatic cancer. Pancreatology. 2020;20: 1502–1510.

57. Zhang G, Yang L, Kim GS, Ryan K, Lu S, O” Donnell Rk, et al. Critical role of sphingosine-1-phosphate receptor 2 (S1PR2) in acute vascular inflammation. Blood. 2013;122: 443–455.

58. Green JA, Cyster JG. S1PR2 links germinal center confinement and growth regulation. Immunol Rev. 2012;247: 36–51.

59. Skoura A, Michaud J, Im D-S, Thangada S, Xiong Y, Smith JD, et al. Sphingosine-1-phosphate receptor-2 function in myeloid cells regulates vascular inflammation and atherosclerosis. Arterioscler Thromb Vasc Biol. 2011;31: 81–85.

60. Grimm M, Tischner D, Troidl K, Albarrán Juárez J, Sivaraj KK, Ferreirós Bouzas N, et al. S1P2/G12/13 Signaling Negatively Regulates Macrophage Activation and Indirectly Shapes the Atheroprotective B1-Cell Population. Arterioscler Thromb Vasc Biol. 2016;36: 37–48.

61. Winkler MS, Nierhaus A, Poppe A, Greiwe G, Gräler MH, Daum G. Sphingosine-1-Phosphate: A Potential Biomarker and Therapeutic Target for Endothelial Dysfunction and Sepsis? Shock. 2017;47: 666–672.

62. Xiong Y, Piao W, Brinkman CC, Li L, Kulinski JM, Olivera A, et al. CD4 T cell sphingosine 1-phosphate receptor (S1PR)1 and S1PR4 and endothelial S1PR2 regulate afferent lymphatic migration. Sci Immunol. 2019;4. doi:10.1126/sciimmunol.aav1263

63. Vastrad B, Vastrad C, Tengli A. Bioinformatics analyses of significant genes, related pathways, and candidate diagnostic biomarkers and molecular targets in SARS-CoV-2/COVID-19. Gene Rep. 2020;21: 100956.

64. Abdolrahimzadeh Fard H, Borazjani R, Sabetian G, Shayan Z, Boland Parvaz S, Abbassi HR, et al. Establishment of a novel triage system for SARS-CoV-2 among trauma victims in trauma centers with limited facilities. Trauma Surg Acute Care Open. 2021;6: e000726.

65. Young BC, Eyre DW, Jeffery K. Use of lateral flow devices allows rapid triage of patients with SARS-CoV-2 on admission to hospital. The Journal of infection. 2021. pp. 276–316.

66. Barnacle JR, Houston H, Baltas I, Takata J, Kavallieros K, Vaughan N, et al. Diagnostic accuracy of the Abbott ID NOW SARS-CoV-2 rapid test for the triage of acute medical admissions. J Hosp Infect. 2022;123: 92–99.

